# Variations in secondary health resource utilisation post Clozapine initiation

**DOI:** 10.1101/2020.05.16.20103929

**Authors:** Prasanna N. de Silva

**Affiliations:** Cumbria, Northumberland, Tyne and Wear NHS Foundation Trust, Monkwearmouth Hospital, Newcastle Road, Sunderland SR1 2AT

**Keywords:** psychosis, health economics

## Abstract

This study looks at secondary care utilisation metrics as an effectiveness indicator of Clozapine initiation in 89 patients over the following year compared to the year prior to initiation. It was found that there was an overall reduction of outpatient use and crisis activity, with a marginal drop in bed usage. Medical outpatient use increased, as did physical health monitoring. Overall, compared with costs over the year preceding Clozapine initiation, an estimated saving per patient of £1800 was estimated over the first year post initiation, increasing to £3100 per patient if those with premorbid dissocial and substance misuse traits were excluded, raising the issue of better treatment targeting.

## Introduction

Clozapine remains the treatment of choice for resistant psychosis (Kane et.al, 2001)) with evidence of improvement around 55% (Raguraman et.al, 2005). Predictors of good outcome has been described previously (Lieberman et.al, 1994), alongside health economic outcome data in North America involving patients with Schizophrenia (Meltzer et.al 1993, Rosenheck et.al. 1999). However, a National Health Service (NHS) Trust wide study of objective metrics such as bed days, psychiatric / medical outpatient episodes, physical health monitoring and crisis team contacts have not been carried out. It is also unknown if specific sub groups, such as patients with premorbid dissocial and substance use traits have outcomes similar to those without such comorbidities. Furthermore, correlation between objective metrics and clinician opinions on effectiveness has not been analysed.

## Objectives of Cloritos

The objectives of this study were agreed beforehand with the Trust Research and Development (R&D) service and the executive medical director. The author (PDS) was selected on not having any relationship with the Clozapine initiation service. The study was under the heading of a service evaluation, requiring the consent to use trust wide electronic data through the Caldecott Guardian (also the executive medical director). The author obtained hypothecated costs of bed days, outpatient and crisis team contacts via the Director of Finance.

## The objectives of the study were

1. To confirm the overall effectiveness figure (as a %) as documented by the responsible consultant at 1 year post clozapine initiation, and compare with that provided by previous studies;
2. To investigate if the clinician observed effectiveness rate was different between patients with or without comorbidity
3. To compare bed days, psychiatric outpatient / crisis team contacts, medical outpatient contacts and physical health surveillance contacts 12 month preceding clozapine initiations versus 12 months post initiation.
4. To use hypothecated costing to describe the effect size of any secondary healthcare costs
5. To ascertain the extent of additional psychotropics (such as other atypical antipsychotics, benzodiazepines, Mood Stabilisers and Antidepressants) used with Clozapine at the end of 1 year.

## Inclusion and exclusion criteria

1. Adults with functional psychoses between 18 – 65, both sexes included. Those with organic psychosis such as traumatic brain Injury, epilepsy and Parkinson’s psychosis were excluded
2. Note taken of pre morbid Dissocial, Borderline traits and Substance abuse, not excluded but analysed separately (as requested by the consultant body)
3. Clozapine initiated between 2010 and 2018 (data integrity assured on Rio database)

## Hypotheses

Clozapine initiation reduced bed day, outpatient and crisis team utilisation over the next year, with equivalent reductions in cost of healthcare for secondary mental health services.

## Method

On ethics approval, consent was obtained via the R&D department despite data collection not entailing patient contacts. Furthermore, approval was sought from the Trust Caldecott guardian to access patient records. Thereafter the author accessed via the Trust pharmacy services the names and Rio numbers of patients attending the 3 Clozapine clinics within the Trust (North, Central and South business units).

The author systematically reviewed the documentation letters, identifying the initial presentation, diagnoses (including comorbidities), date of initiation on Clozapine and ascertained bed days (number of months corrected down), outpatient, crisis team and physical healthcare contacts. Evidence of other psychotropics alongside Clozapine was noted at the end of the 1 year period on Clozapine. Clinician observed outcome was noted at the end of 1 year on Clozapine by examining the most recent GP letter around 1 year in to Clozapine initiation.

Hypothecated costs were estimated as £12,000 bed cost per month (a month calculated as 30 days), psychiatric outpatient appointment cost £ 300 each, medical outpatient appointment cost £ 400, crisis team episode cost £200 and physical healthcare episode £ 100. It was agreed that the annual cost of Clozapine prescriptions was £500 per patient, taking in to account the use of liquid Clozapine used in some ward settings on initiation, which incurs a higher cost.

## Results

### a) Clinicians judgements

Overall, 89 subjects were included in the study; 55 males and 34 female patients between the ages of 23 and 65 years. Regards main diagnosis, 62 were diagnosed to have Schizophrenia (including 3 people with additional Learning Disability), 9 with Schizoaffective disorder, 4 with Delusional Disorder, 6 with Bipolar Affective Disorder and 8 with Emotionally Unstable Personality Disorder with brief psychoses. Of the 89, 14 patients were noted to be abusing drugs (cannabis, amphetamine, ecstasy) before and during Clozapine use, with a further 5 patients using alcohol to excess.

According to their treating consultant psychiatrist, 48 / 89 (54%) responded to Clozapine at the end of 1 year post initiation. Partial or non-responders were characterised by anxiety / social avoidance, continuing substance use (including alcohol) and continuing hallucinations. Patients with premorbid dissocial and substance abuse traits (often associated with forensic contact) did not show any reduction of bed utilisation, but the treating psychiatrists were largely satisfied by their clinical improvement.

### b) Healthcare utilisation

Regards service utilisation, overall there was a marginal reduction of bed day use from a total of 316 to 298 months, with all of the reduction accounted for by ‘responders’ (as identified by their treating consultants). Outpatient use dropped from 266 to 231 episodes (a 13% reduction), and crisis team episodes (usually lasting 1–2 weeks) also fell from 32 to 15 (a 50% reduction).

There was a rise in physical health care appointments; 62 to 112 (an 81% increase), along with a rise of medical outpatient contacts; 58 to 83 (a 30% rise, mainly involving cardiac, diabetic and neurology services). The ‘non responders’ and those with dissocial and substance use premorbid traits had marginally greater use of medical outpatient services.

### c) Hypothecated financial costs and savings

In terms of financial costs, overall Clozapine utilisation produced a cost saving of £ 168,800 for the 89 patients (responders and non-responders), a hypothecated saving of £1,896 per patient. On bed use there was an estimated cost reduction of £ 216,000, on psychiatric outpatient use a total reduction of £7,000, with Crisis team use suggesting a reduction of £3,400 alongside cost rises in medical outpatient use (£ 7,500) and physical healthcare episodes (£ 5,600). The overall cost of Clozapine prescription (including level testing) was estimated to be £44,500. If patients with comorbid dissocial and substance abuse traits were excluded, the ‘pure’ psychosis patients given Clozapine yielded a saving of £199,800; a saving per patient over the first year of £3,171; mainly due to less bed usage post Clozapine. This compared to a cost increase of £19,200 for comorbid patients (£738 per comorbid patient).

## Discussion

1. On weaknesses, this was a retrospective study covering 10 years of Clozapine initiation; clinical practice could have changed during this period regards selection for Clozapine initiation. Furthermore, the number of patients was relatively small (89), perhaps a multi Trust study would provide conclusive findings. The financial estimates pertain to a single NHS Trust, and might not be representative. Furthermore, it was difficult to maintain blinding when counting activity pre and post Clozapine.
2. To the author’s knowledge, this is the first NHS Trust wide naturalistic study examining health utilisation metrics (bed days, outpatient and crisis team episodes) over 1 year following Clozapine initiation involving the full range of diagnostic groups (including people with learning disability and co-morbid forensic issues). There are other studies modelling costs of Clozapine use; mainly North American data specifically studying patients with a diagnosis of schizophrenia (Meltzer et.al, 1993) and Bipolar disorder (Nielsen et.al, 2012). These studies showed cost savings in terms of reduced admissions and overall bed use 2 years post clozapine initiation. NHS studies looking at patients with borderline personality Disorder (Rohde et.al, 2017) and learning disability (Rohde et.al, 2018) initiated on Clozapine showed similar reductions over 2 years. However, community costs and savings including pharmacy costs have not been described.
3. This study demonstrates overall cost savings being evident even within the first year; a hypothecated £1,900 saving per patient over the year (increasing to £3,100 over the year if patients with dissocial / substance abuse features are excluded). Hypothecated costs in this study could be viewed as underestimates; this was purposefully determined to provide conservative estimates. The Crisis service is a statutory requirement, and carry a further fixed cost which was not taken in to account.
4. Clearly, the finding of marginal reduction of bed usage is disappointing. There was evidence that patients continuing to misuse substances did not appear to respond fully to Clozapine confirmed by no reduction of bed usage. Perhaps, substance use was linked to phobic anxiety (also a feature of non-responders). It is difficult to comment on continuing and prolonged bed use among patients who presented with dissocial personality traits and criminal activity prior to developing psychotic symptoms (such as hallucinations). It is likely that risk mitigation procedures in forensic settings could cloud the potential benefit of Clozapine initiation. There is also a possibility that diagnosing possible schizophrenia in these patients might be hazardous among these patients (10 in this study) with pre psychotic dissocial traits; also prone to drug misuse despite their continuing inpatient status.
5. There were a small number of patients receiving other psychotropics in addition to Clozapine, mainly atypical dopamine blocking antipsychotics (10), mood stabilisers (10) and benzodiazepines (6). Patients on additional atypical antipsychotics (but not mood stabilisers and benzodiazepines) showed improvement according to their treating consultant psychiatrists, suggesting potential benefit in Clozapine augmentation with selective Dopamine blockers such as Amisulpiride (as was the case in this study). This has been commented on in other studies (Porcelli et.al, 2012).
6. This sample was too small to detect any clear reduction of self-harm attempts with only 6 patients presenting with suicidality prior to Clozapine initiation. Certainly there was no evidence of increased suicidality post Clozapine initiation, and patients with suicidal ideation (3) did not describe (or demonstrate) suicidality. Interrogation of multiple electronic databases of NHS mental health trusts could answer this important question, including looking at inflammatory markers as a predictor of suicidality (Brundin et.al 2015).

### Service developments

1. It is hoped that to reduce costs, ward staff will be encouraged to reduce the use of liquid clozapine (6 times the cost of oral Clozapine).
2. Stricter management of continuing substance use in in patient settings (even if it means use of Methadone on prescription) could be beneficial clinically and reduce costs.
3. A case has been made for mobile Clozapine testing over the weekends to increase concordance and reduce bed days.
4. The Trust is currently upgrading its electronic database to produce mandatory alerts if a patients are on Clozapine, Lithium and depot antipsychotic. This would significantly improve safety in terms of potential drug interactions and toxicity.

## Conclusion

1. The method employed in this study could be used to assess overall cost utilisation for other drugs such as mood stabilisers, stimulants and memory enhancers.
2. On Clozapine specifically, this drug remains the main option for resistant psychosis; this study showed similar improvement rates to previous studies; around 55%.
3. Whether clozapine initiation should be limited to potential responders is worth discussing further; as there are ethical issues to keep in mind. However, it is a difficult decision to discontinue Clozapine clinically due to the risk of rebound psychosis with the resultant risk of escalation in metabolic and cardiac disease.

## Data Availability

Data availability statement – Data template / bank provided as part of submission

## References

1. Kane, J.M., Marder, S.R., Schooler, N.R. et.al. Clozapine and Haloperidol in moderately refractory schizophrenia; a 6 month randomised and double blind comparison. Arch. Gen. psych. 2001, Vol 58: 965 – 972

2. Raguraman, J., Sagar, K.J.V., Chandrasekaran, R. effectiveness of Clozapine in treatment resistant schizophrenia. Indian Journal of Psychiatry. 2005, Vol 47 (2): 102 – 105

3. Lieberman, J.A., Safferman, S.A., Pollack, S. et.al. Clinical effects of Clozapine in chronic schizophrenia: response to treatment and predictors of outcome. American Journal of Psychiatry. 1994, Vol. 151 (12): 1744 – 1752

4. Meltzer, H., Cola, p., Way, L. et.al. Cost effectiveness of Clozapine in neuroleptic resistant schizophrenia. American Journal of Psychiatry. 1993, Vol 150 (11): 1630–1638

5. Rosenheck, R., Cramer, J., Allan, e. et.al. Cost effectiveness of Clozapine in patients with high and low levels of hospital use. Archives of General Psychiatry. 1999, Vol 56 (6): 565 – 572

6. Rohde, C., Polcwiartek, C. et.al. Real world effectiveness of Clozapine for Borderline Personality Disorder: Results from a 2 year mirror image study. Journal of Personality Disorders. 2017, Vol 31: 1 – 15

7. Rohde, C., Hilker, R., et.al. Real world effectiveness of Clozapine for intellectual disability: Results from a mirror image and reverse mirror image study. Journal of Psychopharmacology. 2018, Vol 32 (11): 1197–1203

8. Nielsen, J., Kane, J.M., Cornell, C. Real world effectiveness of Clozapine in patients with Bipolar disorder: Results of a mirror image study. Bipolar disorders. 2012, Vol 14: 863 – 869

9. Porcelli, S., Balzarro, B., Serretti, A. Clozapine resistance: augmentation strategies. European Neuropsychopharmacology. 2012, Vol 22 (3): 165 – 182

10. Brundin, L., Erhardt, S., Bryleva, E.Y. et.al. The role of inflammation in suicidal behaviour. Acta. Psychiatr. Scand. 2015, Vol 132 (3): 192 – 203

